# TRENDS, IN-HOSPITAL OUTCOMES, AND INDEPENDENT PREDICTORS OF ACUTE KIDNEY INJURY IN PATIENTS ADMITTED FOR MYOCARDIAL INFARCTION WITH PERCUTANEOUS CORONARY INTERVENTION: An insight from the National Inpatient Sample Database

**DOI:** 10.1101/2023.07.14.23292693

**Authors:** Akanimo Antia, Daniel Ubokudom, Olanrewaju Adabale, Ovie Okorare, Emmanuel Daniel, Endurance Evbayekha, Chinwendu Angel Onuegbu, Kenneth Ong

**Author notes:** Corresponding author: Akanimo Antia, MD Lincoln Medical Center 234 E 149th Street Bronx, New York. Conflicts of Interest: All authors declare no conflict of interests.

## Abstract

**Background:** Acute kidney injury (AKI) is an important risk factor associated with adverse outcomes in cardiovascular illnesses, more importantly, myocardial infarction (MI). This study describes the Trends, in-hospital outcomes, and independent predictors of Acute Kidney Injury (AKI) in patients admitted for Myocardial infarction with Percutaneous Coronary Intervention (PCI).

**Methods:** This retrospective study used patient records from the 2016-2020 National In-patient Database (NIS). We identified patients who were admitted for the management of an MI who had a PCI procedure and an AKI and evaluated their associated socio-demographic and comorbid factors using International Classification of Diseases-10 (ICD-10) codes. The chi-square test was used to compare baseline characteristics between our populations with and without AKI and outcomes and multivariate logistic regression to identify independent predictors of AKI.

**Results:** There were 1,551,630 patients admitted for an MI and PCI, with 15% having an AKI. We observed that our population with AKI were older on admission and were more likely to be whites than blacks. A higher percentage were males. Our subpopulation was likely to have heart failure, atrial fibrillation, coronary artery disease, obesity, CKD and Charlson comorbidity index ≥ 3. A diagnosis of AKI was associated with higher in-hospital mortality rates [adjusted odds ratio (aOR): 2.84, CI: 2.7–3.02, p<0.001], longer mean length of stay (LOS) and higher hospital costs. We noted an increasing trend in the percentage of patients who had an AKI, from about 13.5% in 2016 to 16.5% in 2020.

**Conclusion:** Acute Kidney Injury is strongly associated with worse hospital outcomes in patients admitted for MI and PCI, with higher mortality rates, a longer mean length of stay, and a higher hospitalization cost. A more concise look at preventive measures is recommended to minimize these outcomes.

## INTRODUCTION

Acute Kidney Injury (AKI) following percutaneous coronary intervention (PCI) is a common complication occurring in > 15% of STEMI patients who undergo primary PCI ^1, 2^. The use of more contrast agents during the procedure has been linked to a higher incidence of AKI after primary PCI versus elective PCI. ^3, 4^. Emergent primary percutaneous coronary intervention (primary PCI) remains the gold-standard ST-segment elevation myocardial infarction (STEMI) treatment. Recent studies have shown improved outcomes and prognosis for patients with STEMI, with one-year mortality at 11% ^5^.

AKI can occur in patients with both pre-existing impaired renal function and in patients with normal baseline renal function ^6^. In general, acute myocardial infarction (AMI) is one of the critical conditions that can trigger AKI. The most common etiologies stem from hemodynamic instability causing renal hypoperfusion, radiocontrast toxicity, and athero-embolism. About 5% of patients undergoing PCI experience a transient increase in the plasma creatinine value of more than 1.0 mg/dL because of contrast use. The risk is greatest in patients with a history of diabetes mellitus and moderate to severe impaired renal function.

In the past decades, the morbidity of AKI has increased from 3/1000 to 5/1000 in the United States ^7^. More so, acute myocardial infarction (AMI) complicated by cardiogenic shock may increase the incidence of AKI by more than 50% ^8^. Among patients with AMI, those with AKI had a 20- to 40-fold higher mortality rate in comparison to those without AKI. Patients with AKI also have prolonged hospital stays, increased hospital charges, and more long-term complications, including recurrent AMI, heart failure, chronic kidney disease progression, and long-term mortality ^8, 9^.

The aim of our retrospective study is to describe the trends, in-hospital outcomes, and independent predictors of acute kidney injury in patients admitted for myocardial infarction with percutaneous coronary intervention.

## METHODS

This study is reported following the strengthening of the reporting of observational studies in Epidemiology (STROBE) reporting guidelines.

### Study Design

#### Data Source

In this retrospective study, we analyzed hospitalizations between January 1^st^, 2016, and December 31st, 2020, from the National Inpatient Sample (NIS) in the United States (US). The NIS was created and is maintained by the Agency for Healthcare Research and Quality and is the largest publicly available all-payer in-patient database in the United States of America (US). It was designed as a stratified probability sample representing all non-federal acute care hospitals nationwide. Hospitals are stratified according to ownership/control, bed size, teaching status, urban/rural location, and geographic region.

A multistage 20% probability sample of all hospitals within each stratum is then collected. All discharges from these hospitals are recorded and then weighted to ensure they are nationally representative. Data from 47 statewide data organizations (46 States plus the District of Columbia) encompassing more than 97% of the US population is included in the NIS 2016 – 2020 sampling frame. As many as 30 discharge diagnoses for each hospitalization were recorded using the International Classification of Diseases, Tenth Revision, Clinical Modification (ICD-10) in NIS 2016, and 40 discharge diagnoses and 25 procedures were coded in the NIS 2020 database. In the NIS, diagnoses are divided into principal and secondary. A principal diagnosis was the main ICD-10 code for hospitalization. Secondary diagnoses were any ICD-10 code other than the principal diagnosis.

Since all patient data in NIS are de-identified and publicly available, we waived the institutional review board approval.

#### Inclusion Criteria and study variables

The study population consisted of all in-patient hospitalizations with a primary diagnosis or secondary diagnosis of <u>Acute Kidney Injury (AKI) during admission for myocardial infarction and a Coronary percutaneous intervention</u> recorded between 2016 and 2020 following the exclusion of patients below 18 years. The diagnosis was identified using ICD 10 codes recommended by the American college of and the American Association of Cardiology Respectively.

Demographic Study variables, including Age, race, Median household income, Primary insurance, and co-morbidities (computed from the Charlson comorbidities index), were identified as variables already present in the data set. Other variables were identified using The American ICD10-CM (diagnosis) medical billing codes identified from the review of other nationwide studies on cardiovascular diseases (**Table 1**). The ICD-10 codes used to identify our principal diagnoses, secondary diagnoses, and comorbidities are listed in the supplementary Table 1

**Table 1.** Socio-Demographic and Co-morbid Differences in baseline characteristics of patients admitted for MI and PCI with and without AKI.

| Variables | Myocardial Infarction and Percutaneous Intervention<br>N= 1, 551, 630<br>(%) |  | P-values |
| --- | --- | --- | --- |
|  | AKI<br>N= 232,180 (15%) | No AKI<br>N= 1, 319,450<br>(85%) |  |
| **PATIENT CHARACTERISTICS** |  |  |  |
| Age, years, mean ±SE | 68.9 ± 0.1 | 63.5 ± 0.03 | <0.0001 |
| **SEX** |  |  | <0.001 |
| male | 67.5 | 68.3) |  |
| Female | 32.5 | 31.7 |  |
| **RACE** |  |  | <0.0001 |
| White | 71.3 | 76.54 |  |
| Black | 12.21 | 8.97 |  |
| Hispanic | 9.4 | 8.1 |  |
| Asian | 3.1 | 2.6 |  |
| Native American | 0.6 | 0.7 |  |
| Other | 0.3 | 0.3 |  |
| **CHARLSON COMORBIDITY**<br>**INDEX SCORE** |  |  | <0.0001 |
| 1 | 9.6 | 34.2 |  |
| 2 | 16.1 | 30.2 |  |
| ≥3 | 74.3 | 35.6 |  |
| **COMORBIDITIES** |  |  |  |
| Congestive Heart Failure | 53.8 | 21.1 | <0.0001 |
| Atrial Fibrillation | 24.61 | 12.04 | <0.0001 |
| CKD | 49.2 | 11.7 | <0.0001 |
| Anemia | 34.4 | 10.8 | <0.0001 |
| COPD | 18.5 | 12.9 | <0.0001 |
| Obesity | 21.7 | 19.9 | <0.0001 |
| History of CABG | 9.0 | 7.14 | <0.0001 |
| Carotid Artery Disease | 2.5 | 1.4 | <0.0001 |
| Dyslipidemia | 67.3 | 71.6 | <0.0001 |
| Smoking | 26.8 | 27.6 | 0.0018 |
| Dialysis Dependent | 1.3 | 2.5 | <0.0001 |
| Hypertension | 23.4 | 53.6 | <0.0001 |
| Diabetes | 12.0 | 17.9 | <0.0001 |

#### Outcomes

We compared the sociodemographic differences in the population with and without AKI as highlighted in **Table 1**. We further analyzed the trend in the incidence of AKI and outcomes inclusive of (cardiogenic shock, cardiac arrest, and in-hospital mortality) across the years, from January 1^st^, 2016, to December 31^st^, 2020.

#### Data Analysis

Our data was analyzed using STATA, version 17 (Stata Corp, Texas, USA). We performed all analyses using weighted samples for national estimates following the Healthcare Cost and Utilization Project (HCUP) regulations for using NIS databases. Based on descriptive analysis of our data, continuous variables were presented with mean, and standard deviation and the differences were tested using a T-test. Categorical variables were presented with numbers (percentages) and compared with the chi-square test. Trends in incidence, mortality, and complications of AKI in our population were accounted for using conditional marginal effects of the variable “YEARS.” Outcomes were analyzed to obtain adjusted Odds ratios (AOR) using multiple logistic regression and linear regression models to account for potential confounders. The P-values considered significant in the multivariate analysis were two-sided, with < 0.05 as the threshold for statistical significance.

## RESULTS

There were 1,551,630 patients admitted for an MI and PCI, with about 232,180 (15%) having an AKI compared to 1,319,450 (85%) without AKI.

Our patients with AKI were notedly older with a mean age of 68.9 years as compared to those without AKI with a male predominance in both groups.

The white race (75.8%) was more represented than blacks (9.5%) but when isolating each race, AKI was more common in the blacks (12.2% vs 8.97%) than in whites (71.3% vs. 76.5%) (p<0.0001).

Our subpopulation was likely to have heart failure (53.8% vs. 21%, p <0.0001), atrial fibrillation (24.6% vs. 12%, p>0.0001), coronary artery disease (2.5% vs. 1.44%, p<0.0001), COPD (18% vs. 12.5%, p>0.0001), history of CABG (9.6% vs. 7.14%, p<0.0001), obesity (21.7% vs. 19.9%, p<0.0001), CKD (49.2% vs. 11.73%, p<0.0001), Charlson comorbidity index ≥ 3 (74.3% vs. 35.6%, p<0.0001) and anemia (34.4% vs. 10.9%, p<0.0001). They were also more likely to be obese.

Our population without a history of AKI had higher prevalence of Dyslipidemia (71.6% vs 67.3%), Nicotine use (27.6% vs 26.8%), Dialysis dependence (2.5% vs 1.3%) and Diabetes (17.9% vs 12%)

The independent Predictors of AKI in the study population include chronic kidney disease (aOR: 5.3, CI: 5.1 – 5.5, p=<0.001), cardiogenic shock (aOR: 4.9, CI: 4.7 – 5.0, p<0.001), cardiac arrest (aOR: 3.1, CI: 2.9 – 3.3, p<0.001), Charlson comorbidity index of ≥ 3 (aOR: 2.3, CI 2.2 – 2.4, p<0.001) and Heart Failure (aOR:2.1, CI 2.0 – 2.1, p<0.001)

A diagnosis of AKI was associated with higher in-hospital mortality rates [adjusted odds ratio (aOR): 2.84, CI: 2.7–3.02, p<0.001], longer mean Length of Stay (7.1 days vs. 3.0 days, p<0.001) and higher hospital costs ($183,785.8 vs. $101,291.2, p<0.0001).

We noted an increasing trend in the percentage of patients who had an AKI, from about 13.5% in 2016 to 16.5% in 2020.

## DISCUSSION

Acute kidney Injury is a serious and fatal complication of Acute Myocardial Infarction. Deterioration of renal status in patients hospitalized for Acute Myocardial infarction could be influenced by many factors, which include patient hemodynamic status, cardiac function, use of contrast agents and drugs, hypertension or Diabetes Mellitus, underlying CKD, and the performance of CABG ^1, 9–12^.

AKI was defined, according to the European Society of Urogenital Radiology as an increase of 25% in serum creatinine from baseline creatinine within 72 hours from contrast media administration ^2^. In the KDIGO criteria, an increase in creatinine was characterized into stages. Stage 1 is an increase of creatinine by 1.5-1.9 from baseline, and stage 3 is a threefold increase from baseline. This was translated into a severity score with presumptions that a 25% increase based on the European Society of Radiology translates to stage 1 KDIGO ^13^.

Risk factors for AKI have been reported in Patients undergoing PCI, such as old age, Diabetic Nephropathy, time from onset of symptoms to Primary PCI, pre-existing renal disease, congestive heart failure, contrast volume, and length of the procedure ^14–18^. In a recent study with 1656 STEMI patients, congestive heart failure and reduced left ejection was identified as individual predictors of AKI ^19^. This is probably due to decreased cardiac output in patients with myocardial infarction, especially in large infarct size and those with anterior myocardial infarction complicated by cardiogenic shock. Patients with acute coronary syndrome (ACS) tend to also have an increased risk of plaque disruption, particularly cholesterol crystals, subsequent systemic embolism and if the renal arterioles are involved, the risk of AKI increases ^20^.

In addition to the above, our study observed cardiac arrest and a Charlson comorbidity index ≥3 to be strong predictors of AKI in patients that have a PCI procedure done (**table 2**). Contrast volume demonstrated a strong correlation with AKI. In earlier studies, higher risks of AKI were reported in patients receiving 153 -378mls in comparison to those with a lower contrast volume use (111-114 mls) ^4, 16, 17^. Contrast nephropathy has been associated with decreased renal blood flow, causing both intrarenal AKI and prerenal AKI.

**Table 2:**
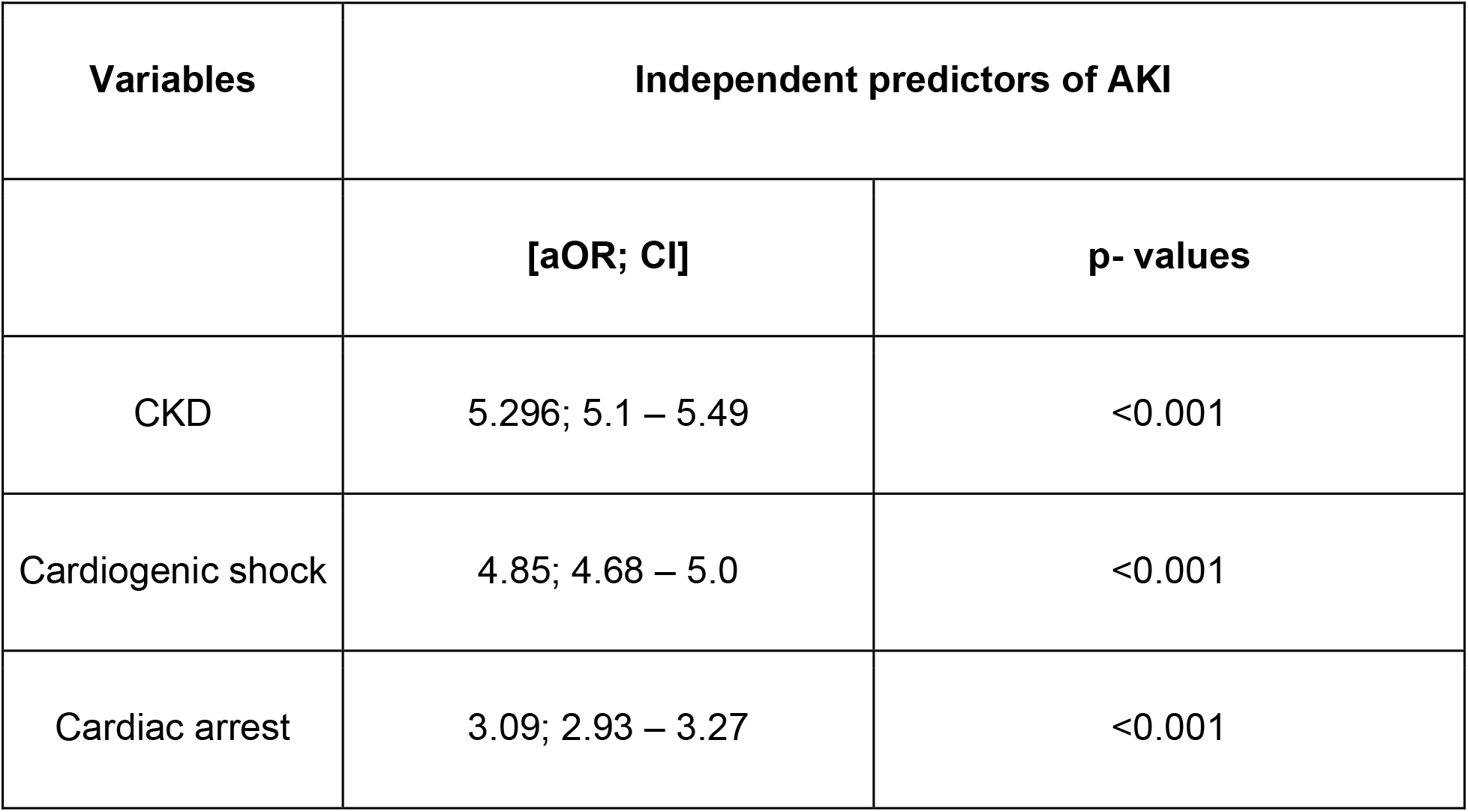
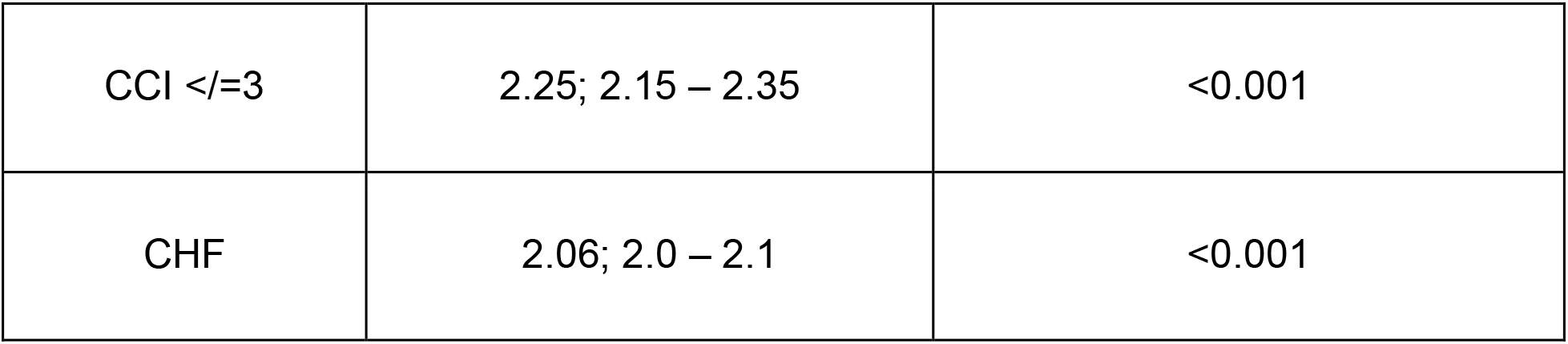
Independent predictors of AKI in patients admitted for MI and a PCI (multivariate analysis)

Medications like furosemide have also been observed as a risk factor for AKI. The use of these loop diuretics has been associated with decreased left ventricular ejection fraction, agitation of the renin-angiotensin system, leading to increased peripheral vascular resistance, renal hypoperfusion, and AKI. Furosemide is commonly used in MI patients presenting with cardiogenic shock and acute pulmonary Edema. Data regarding the use of ACEI and ARBS following early and acute stages of Acute Myocardial Infarction remains controversial. ACEI and ARBs have had great mortality benefits by preventing cardiac remodeling and reducing urine protein. Some studies have shown that the use of ACEI and ARB in patients with insufficient renal perfusion might lead to a decline in glomerular filtration rate, thereby inducing AKI ^21, 22^. Data from a study showed reduced use of anti-RAS agents by doctors due to hyperkalemia and elevated creatinine levels ^23^. A clinical study of 6867 cases showed that the application of anti-RAS therapy in Patients with CKD with ACS could improve 90-day mortality rates^24^. Diabetes Mellitus per se is not an independent risk factor for developing AKI in patients with MI when examined independently. However, when combined with renal insufficiency in patients with MI, the risk increases^25^. Recent studies have shown admission hyperglycemia to be an independent risk factor for AKI following primary PCI 20 ^26^.

Time plays a critical role in acute myocardial Infarction. Early PCI has been shown to limit infarct size, reduce complications of heart failure and decrease risk of AKI^18^.

Our research provided additional evidence that a diagnosis of acute kidney injury is linked to a mortality risk increase of over 50% and longer hospital stays. Furthermore, we noted a substantial increase in the overall hospital expenses incurred by this group of patients (**Table 3**). Efforts have been made to negate the risk of AKI in patients undergoing percutaneous coronary intervention. Some of these measures include reducing contrast volume, Hydrating patients with normal saline infusion and pretreatment with acetylcysteine. Studies by Muller et al. in a large randomized prospective study, showed potential benefits of isotonic saline in reducing AKI especially in unstabilized cardiac patients undergoing PCI ^26^.

**Table 3:** In-hospital outcomes of patients with AKI on admission for MI and PCI.

| OUTCOMES | Myocardial Infarction and Percutaneous Intervention<br>(%), [aOR, CI] |  |
| --- | --- | --- |
|  | AKI | NO AKI |
| Mortality | 11.96 (2.8; 2.67 – 3.01) | 1.72 (ref) |
| Length of Stay | 7.1 (2.24; 2.17 – 2.32) | 3.0 (ref) |
| Total Hospital charge (\$) | 183,785 (61,959; 59451.6 – 64467.9) | 101291 (ref) |

Despite the above preventative measures, our study showed an increasing trend in the rates for AKI in patients receiving PCI procedures (**Figure 1).** This increasing trend could be attributed to the overall rising prevalence of cardiovascular comorbidities in our patient populations.

**Figure 1:**
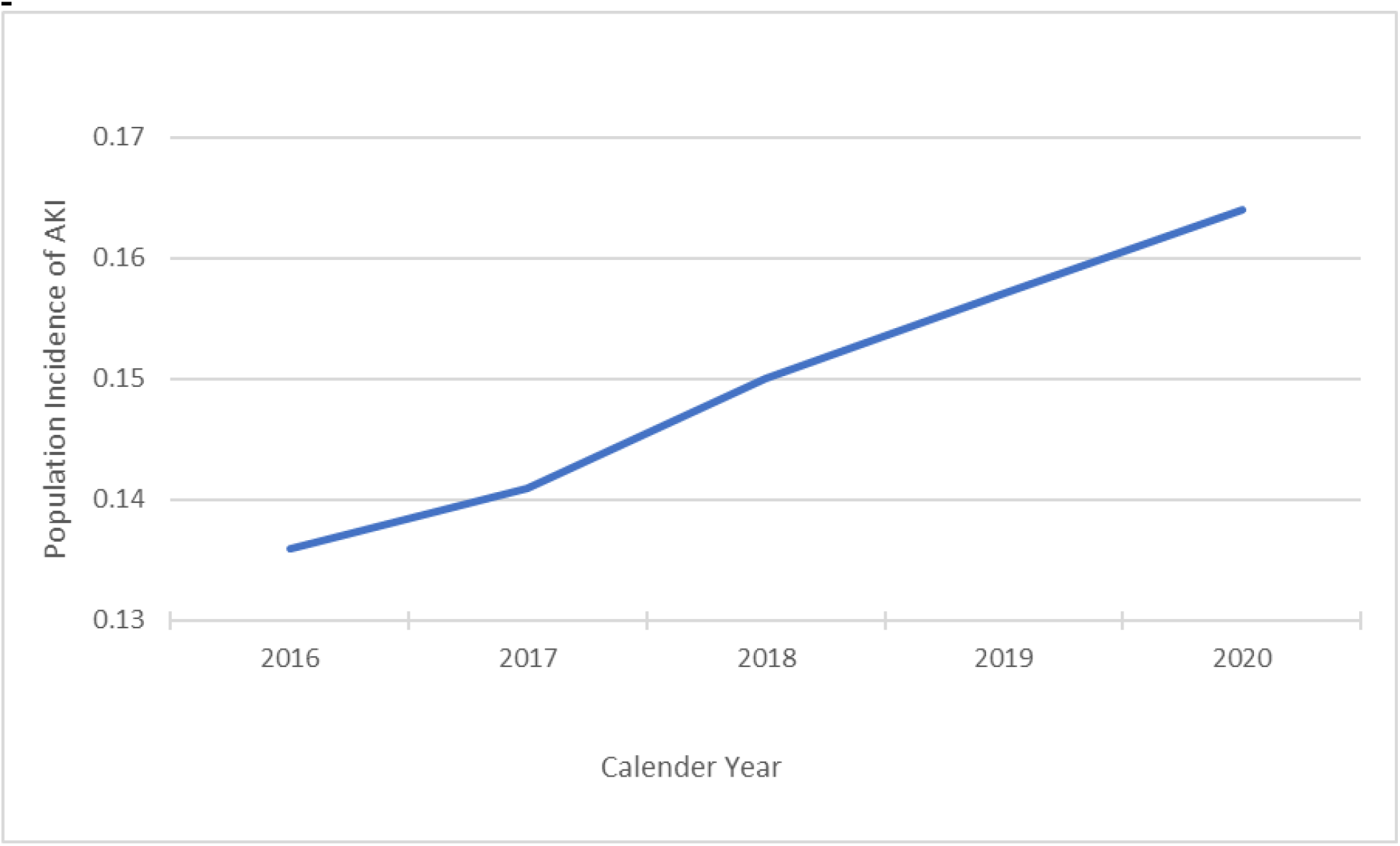
Trends of AKI in patients admitted for MI and a PCI from 2016 - 2020.

In lieu of these trends, we further emphasize, as previously stated, that infusions should be started early and continued long enough to ensure down-trending creatinine levels. Control of Blood glucose in patients presenting with hyperglycemia is necessary to reduce the formation of free radicals which could worsen renal function and myocardial injury. The use of medications such as ACEI and ARBs though beneficial should be individualized keeping in view baseline creatinine. An increase in creatinine of about 25-30% from baseline following use of ACEI and ARBs is acceptable in clinical practice. Intra-aortic balloon pump placement in patients with worsening cardiogenic shock not amenable to diuretics may help improve circulation, reducing the risk of AKI.

Although our study has notable strengths, it also has limitations. Firstly, due to our lack of access to individual-level data, we were unable to assess the consistency of the criteria used to diagnose acute kidney injury, which could potentially result in an over or underestimation of cases. Additionally, as our database only includes in-hospital events, we could not investigate the long-term effects of AKI hospitalization. It is possible that the size of the infarct and patients with lower ejection fractions will have increased risks of AKI but this could not be assessed during this study.

On the other hand, the power of our study lies in its large sample size, which was obtained from a population database, enhancing the credibility of our results.

## CONCLUSION

The burden of acute kidney injury following PCI for acute myocardial infarction is significant, with up to 15% of patients developing AKI following the procedure. It is associated with higher in-hospital mortality, longer mean lengths of stay and higher and higher hospitalization costs. Our retrospective study has shown that independent predictors for AKI following PCI include CKD, cardiogenic shock, cardiac arrest, heart failure, Charlson comorbidity index ≥3, closer attention and monitoring needs to be paid to patients with these risk factors in order to prevent AKI. We also found an increasing tending in the percentage of those that developed AKI from 2016 to 2020 underscoring the need for increased preventative measures.

## Data Availability

The datasets generated and analyzed during the current study are available in the Healthcare Cost and Utilization Project National Data Registry. This Data Use Agreement ('Agreement') governs the disclosure and use of data in the HCUP Nationwide Databases from the Healthcare Cost and Utilization Project (HCUP), which the Agency maintains for Healthcare Research and Quality (AHRQ). Accordingly, HCUP Databases may only be released in 'limited data set' form, as the Privacy Rule defines that term, 45 C.F.R. part 164.514(e). In addition, AHRQ classifies HCUP data as protected health information under the HIPAA Privacy Rule, 45 C.F.R. part 160.103. The datasets generated and analyzed during the current study are not publicly available except for the corresponding author who purchased the data and signed the HCUP Data Use agreement training. Researchers should readily be able to publicly purchase the same databases we did to conduct research.

https://www.hcup-us.ahrq.gov/

https://www.distributor.hcup-us.ahrq.gov/Databases.aspx

## Abbreviations

ACEI: Angiotensin Converting Enzyme inhibitor
ACS: Acute Coronary Syndrome
AKI: Acute Kidney Injury
AMI: Acute Myocardial Injury
aOR: adjusted Odds Ratio
ARB: Angiotensin Receptor blockers
CABG: Coronary Artery Bypass Graft
CCI: Charlson Comorbidity Index
CHF: Congestive Heart Failure
CI: Confidence Interval
CKD: Chronic Kidney Disease
COPD: Chronic Obstructive Pulmonary Disease
HCUP: Healthcare Cost and Utilization Project
KIDGO: Kidney Disease Improving Global Outcomes
PCI: Percutaneous Coronary Intervention
STEMI: ST-Elevation Myocardial Infarction

## Ethics declarations

The study was not submitted for research ethics approval as the activities described were conducted as part of the Nationwide Inpatient Sample Database (NIS), which is part of the family of databases and software tools developed for the Healthcare Cost and Utilization Project (HCUP) and uses de-identified data collected from hospitalized patients. Consent was not obtained, given the use of a de-identified database. All the experiments in our study were under the guidelines and agreement regulations of the Agency Healthcare Research and Quality (AHRQ)

## Authors’ Contributions

Akanimo Antia contributed to the conception and design of the research; Chinwendu Angel Onuegbu contributed to the design of the research; Olanrewaju Adabale contributed to the acquisition and analysis of the data; Ovie Okorare contributed to the interpretation of the data; Daniel Ubokudom, Emmanuel Daniel and Endurance Evbayekha helped draft the manuscript. Kenneth Ong made the final reviews. All authors critically revised the manuscript, agreed to be fully accountable for ensuring the integrity and accuracy of the work, and read and approved the final manuscript.

## Consent for Publication

Not applicable. All data using the National Readmission Database Sample is de-identified.

## Data and Materials Availability

The datasets generated and analyzed during the current study are available in the Healthcare Cost and Utilization Project National Data Registry. This Data Use Agreement (“Agreement”) governs the disclosure and use of data in the HCUP Nationwide Databases from the Healthcare Cost and Utilization Project (HCUP), which the Agency maintains for Healthcare Research and Quality (AHRQ). Accordingly, HCUP Databases may only be released in “limited data set” form, as the Privacy Rule defines that term, 45 C.F.R. § 164.514(e). In addition, AHRQ classifies HCUP data as protected health information under the HIPAA Privacy Rule, 45 C.F.R. § 160.103. The datasets generated and analyzed during the current study are not publicly available except for the corresponding author who purchased the data and signed the HCUP Data Use agreement training. Researchers should readily be able to publicly purchase the same databases we did to conduct research.

---

**Supplemental Tables 1:** International Classification of Diseases, Tenth Edition, Clinical Modification (ICD-10-CM) Codes Used To Identify Comorbidities, and Hospital Outcomes.

| Diagnosis codes (primary and secondary) | ICD-10 codes |
| --- | --- |
| Myocardial Infarction | I21 |
| Percutaneous Coronary Intervention | 0270346, 0272346, 027034Z, 027234Z, 0270356, 0272356, 027035Z, 027235Z, 0270366, 0272366, 027036Z, 027236Z, 027036Z, 0270376, 0272376, 027037Z, 027237Z, 02703D6, 02723D6, 02703DZ, 02723DZ, 02723D6, 0273466, 027146Z, 027346Z, 0271476, 0273476, 027147Z, 027347Z, 02714D6, 02734D6, 02734DZ, 02714E6, 02734E6, 02714EZ, 02734EZ, 02714F6, 02734F6, 02714FZ, 02734FZ, 02714G6, 02734G6, 02714GZ, 02734GZ, 02714Z6, 02734Z6, 02734ZZ, "02703Z6, 02723Z6, 02703ZZ, 02723ZZ, 0270446, 0272446, 027044Z, 02723ZZ, 02723ZZ, 0270446, 0272446, 027044Z, 027244Z, 027244Z, 0270456, 0272456, 027045Z, 027245Z, 0270466, 0272466, 027046Z, 027246Z, 0270476, 0272476, 027047Z, 027247Z, 02704D6, 02724D6, 02704DZ |
| Acute Kidney Injury | N17 |
| **Comorbidities** |  |
| Carotid artery disease | I652 |
| Dyslipidemia | E78 |
| Anemia | D50, D51, D52, D53, D55, D56, D57, D58, D59, D60, D61, D62, D63, D64 |
| Atrial Fibrillation | I48 |
| Chronic Kidney Disease | N18 |
| Obesity | E660, E6601, E6609, E661, E662, E668, E669 |
| History of CABG | Z951 |
| Smoking | Z87891, F17200 |
| Hypertension | I10 |
| Diabetes | E119, E089, E109, E109, E108, E139, E1010 |
| Dialysis Dependent | Z992 |
| COPD | J41, J42, J43, J44 |
| Heart Failure | I50 |
| **Other Variables** |  |
| Cardiogenic Shock | R570 |
| Cardiac Arrest | I462, I469, I468 |
| Abbreviations: MI: Myocardial infarction, CABG: Coronary artery bypass graft, COPD: Chronic Obstructive Pulmonary Disease |  |

